# Can ChatGPT give holistic and accurate patient-centred information to oncology patients? A mixed-methods evaluation with stakeholders

**DOI:** 10.64898/2026.02.02.26345346

**Authors:** Mengxuan Sun, Ehud Reiter, Peter Murchie, Anne E Kiltie, George Ramsay, Lisa Duncan, Rosalind Adam

**Affiliations:** Institute of Applied Health Sciences, School of Medicine, Medical Sciences and Nutrition, University of Aberdeen; School of Natural and Computing Sciences, University of Aberdeen; Rowett Institute, School of Medicine, Medical Sciences and Nutrition, University of Aberdeen and Aberdeen Cancer Centre, University of Aberdeen; Aberdeen Centre for Evaluation, School of Medicine, Medical Sciences and Nutrition, University of Aberdeen

**Keywords:** ChatGPT, MDT report, Cancer Care, LLMs evaluation, Focus groups, stakeholders

## Abstract

**Objective:** More people than ever before are living with cancer. Patient education is a core component of cancer care, and patients are increasingly using large language models (LLMs), such as ChatGPT, for advice. The objectives of this study were to evaluate the ability of ChatGPT to explain specialist cancer care records (multidisciplinary team (MDT) meeting reports) to patients and to understand key stakeholders’ views and opinions about the technology.

**Methods:** Six simulated MDT meeting reports were created by cancer clinicians. MDT reports and 184 realistic patient-centred queries were input into ChatGPT4.0 web version. We conducted a mixed-methods study combining qualitative analysis with exploratory quantitative components to evaluate ChatGPT’s responses. The study consisted of three stages: (1) Clinician sense-checking, (2) Clinical and non-clinical annotation, (3) focus groups (including cancer patients, caregivers, computer scientists, and clinicians).

**Results:** ChatGPT was able to summarise complex oncology information into simpler language, to provide definitions of complex terms and to answer questions about clinical care. However, clinician sense-checking identified problems with accuracy, language and content. In clinician annotation, 92.6% of ChatGPT’s responses were judged problematic. Across all evaluation methods, six recurring themes were identified: accuracy, language, trust, content, personalisation and integration challenges. Patients and clinicians found the summaries and definitions useful; however, the responses were not tailored to the individual patient or to what the report might mean for them.

**Conclusion:** This study highlights current challenges in using LLMs to explain complex cancer diagnoses and treatment records, including inaccurate information, inappropriate language, limited personalisation, AI distrust and challenges in integrating LLMs into clinical workflow. Understanding of the limitations is crucial for clinicians, patients, computer scientists and policy makers. The issues should be addressed before deploying LLMs in clinical settings.

## 1. Introduction

Cancer is the leading cause of death globally, and the burden of cancer is rising around the world. There were 14 million new cases of cancer diagnosed globally in 2012, and this is expected to increase to 22 million new cases by 2030 [1]. Healthcare providers and policymakers expect that digital technologies will improve efficiency in oncology care, support a shift towards self-management and play an essential role in patient education [2, 3].

In recent years, there have been significant advances in investigations and treatments for cancer and the current gold standard of care is that teams of professionals from different disciplines (e.g. radiology, oncology, surgery, pathology) come together to formulate treatment plans in multidisciplinary team meetings [4, 5]. MDT treatment reports are created by most cancer centres globally. They contain multiple sources of data (e.g. from radiology and pathology reports) but are technical, contain dense medical language and are generally not copied to (or accessible to) patients. One potential role for digital technologies is to summarise and explain complex treatment plans and medical reports to patients. Artificial intelligence (AI) can go beyond summarising such reports, to allow patients to ask questions about their treatment plans and have their questions answered. However, the availability of powerful AI tools has outpaced patient-facing research about the scope and implications of such technology.

Large language models (LLMs) are language models trained with supervised machine learning algorithms; they can have trillions of parameters and be trained on terabytes of data [6]. One of the most widely used LLMs is ChatGPT [7], which has over 300 million weekly active users. Inevitably, LLMs are increasingly used by patients and in healthcare settings, including for pre-consultation information and patient education [8]. In principle, patients who have questions about their situation can give their medical reports to LLMs, ask questions and engage in dialogues. However, LLMs have the potential to generate incorrect information which looks believable and trustworthy but could be confusing and misleading [9].

Current research has mainly focused on the accuracy of LLMs, usually by comparing LLM performance against experts [10, 11] and on how clinicians can benefit from LLMs within processes of care [12]. There has been limited exploration of how both clinicians and patients perceive the potential role of LLMs in oncology care and relatively little research into how patients might use and benefit from LLMs.

The aim of this study was to evaluate how ChatGPT performs when asked to summarise and answer questions about complex oncology reports, using simulated MDT reports. An objective was to explore the opinions of patients, clinicians, computer scientists and health service staff regarding the role of LLMs in making complex oncology reports more accessible to patients. In this paper, we use patient-centred to refer to responses that are understandable (low jargon), emotionally appropriate, actionable (next steps), and aligned with the patient’s clinical context and NHS pathways. We use holistic to describe not only biomedical explanation but also support needs (e.g., psychological, practical, and signposting) in a way that does not introduce avoidable distress or misinformation.

## 2. Methods

### 2.1 MDT reports creation

We asked a colorectal surgeon (GR) and a clinical oncologist (AK) to create six simulated MDT reports which accurately mimic reports generated for real colorectal and prostate cancer patients. The clinicians started with real patient reports and then changed key clinical details and removed all identifiable information. The clinicians designed the reports to give a realistic spectrum of cases reflecting the range of patients discussed in a real MDT meeting. All reports were carefully constructed to match real-world MDT documentation in linguistic style, technical content including typical abbreviations, and overall structure. Although synthetic, the full source dataset is included for publication because the reports are highly detailed, are based on the authors’ clinical experiences and could plausibly resemble or be mistaken for real patient records. We used simulated MDT reports to enable reproducible testing across multiple prompts while avoiding risks associated with sharing identifiable or clinically sensitive records with a third-party LLM interface. This design allowed controlled variation in case complexity while maintaining realism in structure and terminology. Most reports were at least one A4 page in length, including four colorectal cancer reports and two prostate cancer reports. The reports included patients of different genders (in the case of colorectal cancer) and ages. Clinical cases ranged in complexity and included recurrence after initial treatment. Supplementary file 1 shows an overview of the MDT reports and their usage in the experiments.

### 2.2 Scenarios and Questions (Prompts) design

To ensure that our questions reflected real-world scenarios, we consulted multiple sources. Questions were adopted directly when appropriate or adapted to fit the reporting context:

- Frequently asked questions (FAQs) from patient resources on widely utilised cancer websites such as Cancer.net [13], the National Cancer Institute [14] and Macmillan Cancer Support [15].
- Cancer online forums, including Prostate Cancer UK [16], Macmillan Cancer Support online forum [17], and Cancer Research UK [18].
- Clinicians’ experiences with common cancer patient questions.
- Realistic potential patient questions based in the clinical experience of the authors.

In total, we collected 184 prompts (questions) for our experiments. To better characterise these prompts, we defined four scenarios: Patient-Explain, Patient-Suggest, Doctor-Explain, and Doctor-Suggest. Descriptions and examples of each scenario are provided in Table 1.

**Table 1.** Experimental scenarios for patients and clinicians for queries to ChatGPT about complex medical reports.

|  | Scenarios | Description | Examples | Source |
| --- | --- | --- | --- | --- |
| 1 | Patient-Explain | The patient has the MDT report and asks ChatGPT to explain it, including the overall content and specific terms. | What is a TRUS Biopsy? Why do I need to do it? | Questions to Ask Your Health Care Team [13] |
| 2 | Patient-Suggest | The patient asks for suggestions based on the information in the MDT report | Approximately how long can I expect to wait from diagnosis to surgery? | Bowel (colon and rectal) cancer forum [17] |
| 3 | Doctor-Explain | The clinician asks ChatGPT to draft an email (which the clinician can edit) to explain the MDT results to a patient. | Write an email to the patient to inform him about his MDT results. | Doctor's expertise |
| 4 | Doctor-Suggest | The clinician asks ChatGPT for recommendations to help patients and outline treatment plans. | What is the patient's prognosis and how should he prepare for the following treatment? | Author generated |

### 2.3 ChatGPT configuration and interaction setup

For each experiment, MDT reports were copied and pasted into the web version of ChatGPT (GPT-4), which was prompted to respond to questions across the scenarios described in Table 1. We conducted the pilot experiments using ChatGPT between October and December 2023. The outputs used in the annotation and focus group exercises were generated between January and February 2024.

As the study involved different user roles and location-specific contexts, each interaction began with a clearly defined scenario. At the start of each dialogue, we specified the user’s role, the referenced MDT report, the user’s requirement, and the geographical setting. To avoid influence from prior interactions, conversational memory was not used and a new dialogue was initiated whenever the MDT report or the user role changed. Further details of the prompting strategy are provided in Supplementary file 2.

### 2.4 Evaluation methods

This study employed a mixed-methods research design to evaluate ChatGPT’s ability to explain complex oncology MDT reports and to explore potential issues in realistic use scenarios. Mixed-methods research integrates qualitative and quantitative approaches, enabling the investigation of research questions that cannot be adequately addressed by one approach alone [19]. Although mixed-methods designs have been increasingly encouraged in healthcare research, prior work suggests they remain underused [20]. In this study, the mixed-methods approach allowed both an initial measurement of key problems and an in-depth interpretation of their significance in practice, thereby enriching the findings and strengthening the credibility of the results. Given that the use of ChatGPT to explain complex clinical reports has not been widely studied, this approach was well suited to identifying real-world challenges and capturing how these outputs are interpreted by different stakeholders.

Evaluation involved three stages: stage one was sense-checking the outputs with clinicians to identify major types of problems in ChatGPT outputs. In stage two, clinicians and non-clinical participants annotated output reports as an exploratory quantitative step, measuring the occurrence of these key issues across model outputs and examining differences in how participants with different professional backgrounds perceived them. In the third stage, three focus groups were conducted with key stakeholders to explore their perspectives in realistic clinical contexts, with an emphasis on understanding the clinical meaning and implications of the issues.

#### 2.4.1 Sense-checking with clinicians

The clinicians who created the MDT reports (AK and GM) and another clinical author (RA) met on Microsoft Teams to review ChatGPT’s outputs and share feedback and opinions about how ChatGPT had handled the tasks of summarising and answering questions about the MDT reports. This initial sense-checking allowed the study team to identify, characterise and categorise errors or other concerns. Insights from this sense-checking process informed the design of the subsequent annotation questionnaires. More examples for sense checking are given in Supplementary file 3.

#### 2.4.2 Non-clinical and clinical annotation

Academic clinicians and non-clinical participants were recruited via the University of Aberdeen email lists. Clinical participants were recruited using purposive, availability-based sampling [21]. Non-clinical participants were recruited via convenience sampling [22] and included researchers without formal cancer expertise. Their research training was considered appropriate for completing the complex annotation tasks.

Two colorectal cancer cases and two prostate cancer cases were chosen for non-clinical participants and three of them (excluding one colorectal case) were provided to clinicians. The selection aimed to balance case diversity across cancer types while ensuring that materials were distributed to multiple annotators, thereby reducing potential bias arising from over-reliance on any single case and allowing exploration of variations in annotation across annotators.

Eight questionnaires were developed, each containing a structured guidance for annotation, an MDT report, and three ChatGPT responses. The participants were asked to read the guide and context, annotate ChatGPT responses, highlight and describe (free text) the problems and suggest any improvements that would make the responses more acceptable to patients. No formal training or calibration was conducted. This was a deliberate design choice to capture natural variation in interpretation, reflecting real-world differences in how patient-facing AI outputs are perceived across roles. Each response was approximately one A4 page long, and each questionnaire took about one hour to annotate. An example is given in Supplementary file 4. After annotating each response, participants were asked to answer the following questions using a 5-point Likert scale (from strongly disagree to strongly agree):

- I think ChatGPT handled the question well (all participants).
- I would accept it if my doctor answered like above (non-clinical participants only).
- I believe the output from ChatGPT meets the standards required in clinical care (clinicians only).

Content analysis was conducted in which [23] problems identified in the sense-checking experiments were quantified by counts during annotation, while free-text comments were collected as qualitative data. To exploratorily assess internal agreement, Likert-scale questions were analysed using Cohen’s Kappa on a 20% data subset, aiming to assess consistency among non-clinical participants and clinicians, as well as differences between them.

#### 2.4.3 Focus groups

Focus groups were conducted to examine and discuss the role of ChatGPT and emerging technologies in cancer care more broadly. The aim was to generate debate and discussion around these technologies and to understand the current perspectives of patients, clinicians, and stakeholders regarding their potential roles and limitations.

Focus groups were conducted during a full day co-design event that examined the role of digital technologies in improving cancer care [24]. Anyone who could communicate in English and who had an interest in digital systems for cancer aftercare was eligible to participate. Professional participants were recruited via mailing lists and snowball sampling (whereby interested participants could share the invitation throughout their networks). Patients were recruited from NHS oncology clinics and through charities and patient/public involvement networks [24]. Participants were divided into three groups, which were purposively mixed to ensure each group had individuals from a range of experiences and backgrounds (i.e., included clinicians, patients, researchers, and professionals).

At the start of the event, all participants were given a one-hour introduction to natural language processing technology by an expert (ER). The session covered the ChatGPT interface, the use case to explain MDT reports, its capability to answer questions and a representative error case. This ensured that participants with no prior knowledge of ChatGPT or similar systems would understand any confusing terminology and feel confident offering opinions about the case studies.

Groups were facilitated by three experienced researchers (authors RA, AK and PM). The facilitators met beforehand to ensure that they were aligned on the core topics for discussion and that the groups would be conducted in a similar fashion.

Three MDT reports were used as case studies during the focus groups. Each group was presented with one case study, which included instructions, reading context, discussion questions and three ChatGPT responses, selected from three scenarios based on the annotation questionnaires. Supplementary file 5 illustrates the prompts and ChatGPT outputs shown to different groups. The participants in the focus groups were shown ChatGPT’s responses but not the MDT reports, as the complexity of MDT reports could be confusing to individuals without a medical background. The guiding questions addressed perceptions of the reports (strengths, weaknesses and areas for improvement), trust in ChatGPT for personal health queries and the potential role of such technologies in cancer care. Focus group discussions, therefore, explored views on ChatGPT outputs derived from MDT reports, key strengths and problems identified, and the opportunities and challenges of implementing LLMs in oncology practice.

Focus groups were audio-recorded and each group discussion lasted 1 hour. Recordings were automatically transcribed into text using Microsoft Word and checked for accuracy. Thematic analysis [25] was used to categorise and identify the key themes under discussion. An inductive, data-driven approach was adopted, with codes developed directly from the transcripts data rather than being pre-determined.

The analysis involved six stages [26]: familiarisation, generating initial codes, searching for the themes, reviewing themes, defining and naming themes and reporting the thematic analysis. Familiarisation with the data was achieved through repeated reading of the transcripts. Line by line coding of all transcripts was performed by MS. NVivo 12 was used to facilitate coding. RA also reviewed the coding to ensure that key concepts were not overlooked. Initial themes were generated by examining relationships between codes and common features in the data. Themes were refined through review for coherence within and distinction between themes and discussion among the authors. Illustrative quotations are reported for each theme to support interpretation, with themes derived from recurrent patterns across the focus group data rather than from isolated excerpts.

### Ethical Considerations

The annotations experiment was approved by the School of Computing Science Ethics Committee, University of Aberdeen. Focus groups were conducted as part of a larger co-design event which was approved by Yorkshire and The Humber—Leeds East Research Ethics Committee, reference 19/YH/0291 and received NHS R&D approval (NHS Grampian). All participants provided written informed consent.

## 3. Results

Overall, six main themes were identified across the three stages of this research. These were: accuracy, language, content, trust, personalisation and integration.

### 3.1 Main issues identified from sense checking with clinicians

Initial sense-checking with clinicians identified problems with accuracy, language and content. Clinicians noted inaccurate responses. These were outputs that contained factual errors, misinterpretations of clinical information or misleading statements, which could cause misunderstandings or create clinical risk.

Language problems included issues with the linguistic style or tone of the response, such as the use of overly technical terminology, inappropriate phrasing (e.g., “Americanised” expressions for UK patients or a tone that “resembled a business transaction” rather than a clinical response) or inappropriate language (e.g., lack of empathy).

Content problems included responses that appeared relevant but failed to adequately address the user’s question or provide actionable information (e.g., too vague to be helpful), thereby limiting their practical value. These problems were elaborated upon and noted more formally in the annotation process.

### 3.2 Non-clinical and clinical annotation

Five academic clinicians and eight non-clinical participants (researchers without any cancer expertise, mainly computing scientists) provided annotations. Annotations highlighted examples of accuracy, language, and content problems. These are summarised in Table 2. Important accuracy problems included a web link that directed patients to an Indonesian gambling website rather than a local cancer support organisation, and summaries that included the wrong numerical value for a blood test result. There were examples of insensitive language, including ChatGPT suggesting to a patient that they should get their “affairs in order” (the implication being that they might die soon). Non-clinical annotators asked for more detailed content, particularly in terms of links to the cancer organisations mentioned in the support information. Content was also noted to be generic and mentioned services that were available in the local area which might not be relevant to the patient’s presentation.

**Table 2:** Examples of problems noted in the lay and clinician annotation exercise

| Type of problem | ChatGPT response | Annotation |
| --- | --- | --- |
| **Accuracy problems** |  |  |
| Unsubstantiated assumptions | "The good news is that we've caught this early" | Non-clinical annotation: *Not mentioned in the report.* |
| Incorrect treatment summary | "Radiation Therapy: Duration: Often 5 days a week for several weeks." | Clinician annotation: *Most of our radiotherapy is 5 days long.* |
| Incorrect web link or reference | http://www.clanhouse.org for the organisation CLAN cancer support, | Clinician annotation: *The webpage for CLAN is incorrect; it redirects to an incorrect site. The actual URL is https://www.clancancersupport.org/* |
| Misreporting of test results | "...since your current PSA is 6.2" | Clinician annotation: *PSA of 6.2 was presenting PSA in the history. Current PSA is 0.050 (post-op).* |
| **Language problems** |  |  |
| Language insensitive | "Legal and Financial Planning: It may be helpful to get your affairs in order, including any legal and financial planning, which can provide peace of mind for you and your family." | Clinician: *This is really poorly phrased. It implies the patient is actively dying and I'd imagine that is how patients would read this sentence to mean* |
| Language insensitive | "I hope this letter finds you well" | Clinician annotation: *This is rather twee and a potentially unwise start to a patient with known malignancy* |
| Lack of empathy | "The recurrence of cancer and its spread to lymph nodes suggest a more challenging situation than the initial diagnosis." | Clinician annotation: *Whilst accurate, I'm not sure I'd use the phrase challenging to explain this type of problem to a patient. It is somewhat confrontational and lacks any empathy* |
| Sentimentality and assumptions about patient experience | "You've been incredibly strong and resilient through your journey with low rectal cancer, initially responding well to the treatments, including the neoadjuvant chemoradiotherapy and the subsequent surveillance. Your courage and strength during | Clinician annotation: *For me, this is over-interpretation. From the data submitted, it is impossible to state how she has coped with this process. Some patients do not cope well and it is too excessive to make this statement* |
|  | these times have been truly admirable.” |  |
| **Content problems** |  |  |
| Lacking detail | “Cancer Specific Support - Macmillan Cancer Support - Cancer Research UK” | Non-clinical annotation: *I would have liked to see contact details next to each organisation that is listed.* |
| Not tailored to the patient | “Grampian has specific services and pain management clinics that can be utilised for symptom control.” | Clinician annotation: *This is unlikely to be an issue for someone with low grade prostate cancer on active surveillance.* |

Table 3 summarises the scale of the problems identified in the sense checking experiments through annotation. Non-clinical participants identified fewer inaccurate responses than clinicians but frequently noted content-related issues, including that the content was too technical, generic or misaligned to the patient’s needs. The most frequently reported issue was the use of medical jargon, which seven non-clinical participants found difficult to understand. Four participants found responses vague and requested more detail, while another four expressed concerns that the responses did not align with clinicians’ procedures and preferred direct involvement with healthcare professionals.

**Table 3.** Non-clinical and clinician annotation measurement for the identified issues. Problems found are the count of annotated comments by different issue categories. Likert ratings report the average of the responses we collected from different participants (3.83 is the average of 23 scenarios through non-clinical annotation, 3.16 is the average of 27 scenarios through clinician annotation). Internal agreement measures the responses of each participant’s group.

|  |  | **Non-clinical annotation** | **Clinician annotation** |
| --- | --- | --- | --- |
| **Number of participants (n)** |  | 8 | 5 |
| **Number of scenarios (n)** |  | 23 | 27 |
| **Problems found (n)** | Accuracy | 4 | 18 |
|  | Language | 16 | 40 |
|  | Content | 12 | 46 |
| **Likert ratings 1-5 (average)** | I think ChatGPT handled the question well. | 3.83 | 3.16 |
|  | I would accept it if my doctor answered like above (non-clinical) | 3.33 | N/A |
|  | I believe the output from ChatGPT meets the standards required in clinical care. (clinician) | N/A | 2.21 |
| **Internal agreement (Cohen's Kappa score)** | I think ChatGPT handled the question well. | -0.1 | 0.28 |
|  | I would accept it if my doctor answered like above (non-clinical) | -0.42 | N/A |
|  | I believe the output from ChatGPT meets the standards required in clinical care. (clinician) | N/A | 0.54 |

Agreement on the Likert-style question shows a considerable variability in non-clinical evaluations, with very low concordance among annotators. Notably, one participant strongly disagreed with the patient-explain response, and two participants strongly disagreed with the doctor-suggest responses.

Clinicians, by contrast, reported problems in 25 of 27 scenarios (92.6%) and were generally more critical than non-clinical participants. They highlighted more issues overall, particularly concerning language and content and were more negative than non-clinical annotators about applying ChatGPT in oncology care. The internal agreement (Cohen’s Kappa) was moderate, suggesting more consistency among clinicians than among non-clinical participants.

The agreement between non-clinical people and clinicians was also assessed. The Cohen’s Kappa score for how well ChatGPT handled the questions is 0.036. For the question of whether the response would be acceptable or meet medical standards (reflecting attitudes towards applying ChatGPT in real-world scenarios), the score is 0.056. These values indicate a low level of agreement between doctors and patients.

We quantified inter-rater agreement using Cohen’s kappa and this approach should be interpreted in relation to the task. The evaluation involved subjective judgements of summary quality, tone and clinical appropriateness, which are known to yield lower agreement across roles. In this study, lower agreement reflects genuine differences in perspective rather than annotation inconsistency and represents an important finding regarding role-dependent interpretations of problematic AI-generated oncology communication.

### 3.3 Focus groups

There were 23 participants, including seven clinicians (from diverse backgrounds including oncology, psychiatry, renal medicine, radiography and general practice), five computer scientists, two health researchers, three National Health Service (NHS) IT experts, five patients who had experienced cancer (one lung cancer, one pancreatic cancer, one colon cancer, one prostate cancer and one unknown cancer) and one caregiver. Some people had multiple roles, for example, some professionals had also experienced cancer or had cared for a loved one who had cancer. We have listed their main role.

The focus group discussions with stakeholders provided a richer perspective, reinforcing earlier concerns and identifying additional themes around trust, personalisation and integration. Trust and accuracy of responses were inter-linked.

### Accuracy and Trust

Clinicians were aware that LLMs could generate inaccurate responses. One NHS IT expert pointed out that the reliability and accuracy of the responses relied on the user putting in appropriate questions/prompts. Both patients and professionals expressed concerns about reliability, risks, and the appropriate use of AI in clinical care. The perception that these risks are likely and not well understood was seen as a major barrier to acceptance.

> *“I recently went to a webinar… they were saying at this point one third of it is still rubbish. I mean when you get to one percent … we’ll be happy… when a third of it is rubbish, you think in health, that’s just not acceptable”.* Consultant clinician

Almost all cancer patients in the focus groups said they would struggle to trust ChatGPT or to use AI-generated suggestions. One caregiver suggested that she would be able to cross-reference and check the information and advice from other online sources.

> *“I would be happy to get this information because I can then Google and find out a bit more about it, so I put that down as a good thing to get that information because you’ve had all these tests and these are the results.”* Caregiver

One patient explained that his trust and confidence in clinicians was tied to their qualifications and experience and it was more difficult for AI to earn this trust.

> *“It’s when I speak to the doctor. I know that the doctor’s been in medical school for seven years and then they’ve got 20 years’ experience or whatever, you know, I get confidence.”* Patient

### Language

Problems with language were common. Both patients and clinicians found that the wording was often too technical, used inappropriate terms, or failed to convey empathy effectively. In the focus group, one ChatGPT summary stated that “the treatment method is neoadjuvant chemotherapy”. A patient noted the complexity of this language:

> *“I think it’s very technical. To me it looks like the report was just designed for a medical expert. There’s (…) nothing to do with patient at all. I don’t feel as a patient that I really get it.”* Patient

Nevertheless, people agreed that the final summary paragraphs of the ChatGPT responses were succinct and useful.

> *“I felt the summary was nice, and in particular, the last paragraph was good.”* Clinician
>
> *“I would agree with him that I felt, having read through that and not being a medical person, I thought I hadn’t a clue, the actual terminology as such. Still, when it got to the bottom, it was almost as if the dialogue changed completely, and it was much more in layman’s terms and it was much more user-friendly to digest. That was good.”* Patient

### Content

Many ChatGPT responses were too general or lacked patient-specific detail and failed to provide the clarity or actionable guidance expected in clinical communication, which could leave patients confused or anxious. For example, one ChatGPT response described recurrence and the need for follow-up, but the timescale of that follow-up was unclear. The patient found the response anxiety-provoking.

“*There’s no mention of time durations, especially there’s talk of recurrence or possible involvement of another lymph node. To me, it’s triggering and it’s a bit anxiety-provoking to think oh my God, it’s not gone away completely, I need to see somebody and it says I do need to see the oncologist and the consultant but when is that going to be. Is that urgent, is it not urgent?”* Patient

### Personalisation

ChatGPT responses often failed to account for individual patient backgrounds or clinical situations, limiting their relevance and usefulness. Both patients and professionals stressed the importance of personalising information to each patient’s needs and context. Participants identified that ChatGPT was able to define a concept (e.g. Gleason staging of a tumour) but there were deficiencies in how this concept was then explained and contextualised to the individual patient. Participants wanted the technology to go beyond offering definitions and explain what the information meant for them. For example, if doctors discussed the Gleason stage with a patient, they would frame the discussions within the context of that individual’s other MDT findings and the implications the stage would have for the suggested management plan.

> *“It tells you, so the reports obviously said the PSA level is 5. Now, the normal range is 4.5, and this patient’s level is between 4.5 and 5.4 and is 5. So, what that means is that ChatGPT is then telling that the patient’s PSA is a marker used to screen for prostate issues; that’s not what the question is. The question is, what does 5 mean? And in this instance, it means you know the disease is stable, I would think.”* Clinician

There was also a concern that giving vague advice or generic instructions could generate more clinical workload.

> *“The final message is to phone your GP. You know that it’s not actually helping. (…) it’s not going to be useful information. It’s very sort of superficial, generic and non-specific, not related to the patient and not particularly useful.”* Clinician

### Integration

Participants had concerns about the difficulties of integrating AI into established healthcare systems. Integration barriers went beyond the performance of ChatGPT and included the need for clinician validation, data privacy concerns and continuous tool refinement.

A senior clinician interested in innovation discussed another clinical service (mental health) that had tried to pilot test using LLMs to summarise clinical records. The clinician had been unable to test or implement any potential solutions within that service due to the approvals required as well as data security and governance concerns.

> *“…we can’t run that in the open AI model because of patient confidential information. So, we wanted to run a small model within our firewalls, but it’s been really, really challenging to even get into that stage, I think, because of the infrastructural challenges plus the information governance challenges that we face.”* Clinician

Similarly, an NHS IT specialist described issues in sharing technical information with patients, and limitations with the infrastructure that allows the health service to communicate with patients.

> *“How do you get it to the patient? And if it’s an electronic format, again with my information governance hat, there’s a lot of information there to provide electronically to a patient, and information governance leads have a little bit of anxiety if it’s not in a letter that’s going through the post.”* NHS IT specialist

## 4. Discussion

### Summary of Findings

Across sense checking with clinicians, non-clinical and clinical annotation and a focus group study, several common issues became apparent in how ChatGPT processed complex clinical information from MDT reports. Six themes were identified: accuracy, language, trust, content, personalisation and integration. These themes illustrate the specific errors and limitations of ChatGPT and the wider challenges of integrating AI-generated outputs into oncology care.

In the annotation exercise, lay participants often focused on language and content issues, such as jargon, vagueness or misalignment with patient needs. Clinicians reported a higher frequency of overall issues and rated ChatGPT’s responses more critically. Agreement within and between groups was generally low, highlighting that there is variability in how different individuals perceive ChatGPT’s outputs.

Focus group discussions enriched these findings by explaining why such problems matter in practice and by surfacing broader concerns about trust, personalisation and integration of AI.

There was general agreement that LLM technology shows promise in summarising complex medical data. However, our research highlights deeper problems with applying ChatGPT in real-world scenarios. Beyond the inherent issues of existing LLMs, we summarised several key points:

1. Digital misdirection could be a significant problem if the error is not so obvious. e.g. redirected to alternative healthcare websites. Participants without a medical background were less able to identify such errors.
2. Different countries have different health care systems, and LLMs sometimes ignore this and generate responses based on a US system, implicitly treating it as globally valid. Perhaps because there is more US material than UK material in ChatGPT training data. Such limitations should be recognised, particularly by computer scientists, as modelling does not always account for cross-national differences.
3. There are varying attitudes towards ChatGPT when explaining complex medical reports. This aligns with the growing research interest in perspectivist approaches [27] and in embracing variation in NLP [28].
4. The empathy demonstrated by ChatGPT does not align with the empathy required in patient communication.
5. ChatGPT does not fully understand how to communicate with patients, and some of its language poses significant risks from a medical perspective. This may be because doctor-patient conversations are rare in ChatGPT training data.
6. Even if the responses are accurate and the language is appropriate, some explanations and recommendations are too general to be helpful to patients.
7. Trust emerged as a major issue. Patients and doctors were reluctant to trust ChatGPT responses unless these had been checked, preferably by clinicians, and some patients did not trust them at all.
8. Integrating this technology into health services poses challenges and would require addressing data privacy and security concerns as well as the quality of accuracy, language and content.

Computer Science techniques such as prompt engineering and fine-tuning could enhance the performance of the language model. Additionally, it is possible to incorporate safety-checking tools to detect issues like spam URLs, but such techniques will reduce the frequency of problems, they may not eliminate them entirely. At a minimum, patient-facing explanations would require explicit mechanisms to flag uncertainty and potential inaccuracies, as well as clear guidance for users on the system’s intended scope and limitations. Any such deployment would also require clinical evaluation, with associated risks made explicit.

Patients emphasised that responses should avoid medical jargon and clearly explain the personal implications and significance of the medical information being provided to them. Furthermore, the entire system must be user-friendly and accessible, especially for elderly cancer patients. Given ongoing concerns about trust, some individuals may be unwilling to engage with AI-based tools; such preferences should be respected. Patients should not be required to interact with AI systems, and explicit consent may be necessary when AI is used in clinical practice.

### Implications for implementation and clinical workflows

Reducing the frequency of problems will make it more realistic to use LLMs to draft patient-facing letters (the Doctor-Explain scenario) which doctors could check and edit. Several doctors said that this could be one of the most useful scenarios for improving efficiency. We note that both doctors and patients wanted doctors to validate ChatGPT responses. The time spent checking and editing ChatGPT responses needs to be realistic for clinicians.

In practice, the minimum quality requirement for such systems is not that their outputs be perfect, but that their use saves time and cognitive effort compared with fully manual writing. Whether this is achieved depends on the clinician, the clinical context, and the user interface [29]. AI content-generation systems for clinicians are often deployed in workflows where either (A) clinicians check and post-edit their output before it is released, or (B) the AI systems generate draft content which clinicians can paste into otherwise manually-written documents [29].

Evidence from real-world clinical evaluation supports this view. Moramarco [30] evaluated a clinical consultation note generation system in live practice and found that clinicians initially ignored the system output, relying entirely on manual note-taking. Over time, however, all clinicians began to incorporate system-generated text into their notes, typically by copying and adapting relevant sections. The finding [30] showed a 9.06% relative decrease in the median times when using the clinical consultation note generating system, and clinicians reported that while it did not save them time, the system was valuable as a tool for double-checking their own notes. Duggan et al [31] examined the use of an ambient scribe tool in outpatient clinical settings to assess its impact on documentation efficiency and clinician workload. Use of the tool was associated with a 20.4% reduction in time spent on clinical notes per appointment, a 9.3% increase in same-day appointment closure, and a 30.0% reduction in after-hours documentation time per workday.

Recent versions of GPT may already be sufficient for some situations, while offering limited or no benefit in others. Their effective integration depends on clearly specified clinical requirements rather than general-purpose deployment. Making the system more helpful will require careful system design that tailors models to specific clinical tasks and data and embeds them within workflows that clearly define the roles and responsibilities of both clinicians and AI systems.

## 5. Context with other research

### The role of ChatGPT in handling complex cancer reports

The advanced natural language capabilities of ChatGPT hold promise in processing complex medical information, such as facilitating the efficiency of doctor-patient communication and supporting clinicians to make treatment decisions. A study reported substantial concordance between ChatGPT-generated management plans and multidisciplinary team (MDT) recommendations on Stage IV, recurrent, synchronous colorectal cancer, which were analysed for adherence to oncological principles using 30 cases [32]. Derton et al. [33] analysed the linguistic features among different demographic groups using 230,325 clinical notes from 5,285 patients treated with radiotherapy from 2007 to 2019. They applied a variational autoencoder topic model to explore social needs in cancer patients’ notes to inform clinicians. Several studies have investigated the use of ChatGPT to assist doctors in multidisciplinary tumour board decision-making [34-37]. Multidisciplinary tumour boards are different specialists who work together closely, sharing clinical decisions specifically in cancer care [38]. These studies generally reported positive performance in selected question categories but consistently noted that model outputs were not equivalent to expert clinical judgement. Three studies [34, 36, 37] evaluated ten cases each, focusing on treatment suggestions and adopted expert-rated concordance, whereas another [35] evaluated 157 cases across preoperative and postoperative scenarios using κ-based concordance analysis. In radiology, ChatGPT has been tested for generating patient-friendly reports and automating lung cancer staging by system performance metrics [39-43], with results suggesting potential value but also variable accuracy. While these studies indicate potential value, they largely focus on accuracy and task performance, leaving the patient-facing clinical interpretation of complex reports underexplored.

### Evaluation of LLMs in medicine

A few studies have reported that ChatGPT can pass medical examinations [44-47], although its responses may vary with prompting [48]. Some evaluations have relied on expert review, with Mehnen et al. [49] examining diagnostic accuracy and Tang et al. [50] assessing medical evidence summarisation. The results [49] show that only 40% of cases were resolved with the first suggestion, and LLMs may produce factually inconsistent summaries and make overly convincing or overly assertive statements, potentially causing harm related to misinformation [50]. Concerns have also been raised about data contamination in LLM [51]. More structured frameworks, such as “AMIE” [52], have assessed broader clinical competencies, reporting generally positive expert and patient feedback. Three quantitative studies were conducted to evaluate the ability of LLMs to respond to cancer questions using private data [53-55]. They have examined safety, consistency, domain coverage and correctness, highlighting strengths in relevance but gaps in completeness. However, the perspectives of patients, clinicians and other relevant stakeholders have not been adequately captured or compared.

### Strength and limitations

Several limitations of our study should be acknowledged. First, recruiting a large number of doctors and patients for annotation posed challenges, limiting the sample size and potentially affecting the generalisability of our findings. In particular, the types and frequency of issues identified may reflect characteristics of the selected cancer pathways rather than the full range of MDT documentation used in clinical practice. Nevertheless, the focus group experiment constituted a relatively large-scale qualitative design, enabling in-depth exploration of key issues across diverse perspectives. Additionally, the demographic characteristics of the focus group participants were not comprehensively collected, which might have introduced bias or limited the representativeness of the results.

Lastly, the main AI tool we applied is the webpage version of ChatGPT4, and new versions are constantly evolving. To check that our findings from ChatGPT4 remain salient, we ran a small, controlled follow-up comparison experiment with ChatGPT4.1 (model gpt-4.1-2025-04-14) as a robustness check. Two MDT questionnaire expert annotation tasks were repeated using identical MDT reports, prompts, evaluation criteria and parameters. The same two clinical authors (AK and GR) independently annotated the ChatGPT-4.1 outputs, following the same procedure used for clinical annotation in the main study. The clinicians were also provided with the corresponding ChatGPT-4 outputs to enable direct, within-rater comparison across model versions. The clinical authors found that the newer version had more problems with accessibility and that there was little attempt to summarise the key information and lots of technical jargon. These findings suggest that challenges in generating patient-centred explanations of complex clinical reports may persist across model updates, underscoring the need for continued, context-specific evaluation rather than assuming that newer versions will automatically resolve such issues.

### Conclusions and recommendations

This study explored ChatGPT’s potential to support the explanation of complex medical reports. This study combined insights from computer science and medicine and engaged both clinicians and patients. The findings highlight the importance of tailoring AI tools to the needs of doctors and patients, ensuring they provide relevant, trustworthy and practical support within clinical workflows. Future research should focus on understanding how AI systems are designed to complement rather than replace existing methods, and on exploring patient–AI interactions to identify effective ways of engaging patients, such as providing examples or encouraging follow-up questions. Addressing key challenges, such as information security, data governance and compliance with healthcare regulations, remains crucial to safe adoption. Advances in prompt engineering, model fine-tuning and in-context learning, which personalise responses based on patients’ prior knowledge or conversations, may further improve communication quality. Overall, a multidisciplinary effort between NLP researchers, healthcare professionals and patients is essential to create AI systems that are both technically robust and meaningfully integrated into clinical practice.

## Data Availability

The data that support the findings of this study are not publicly available due to privacy and ethical restrictions, but may be made available from the corresponding author upon reasonable request.

## Funding

This study was funded as part of a personal fellowship awarded to Dr Rosalind Adam by Chief Scientist Office (Scotland), reference SCAF18/02. Miss Mengxuan Sun also received funding from the Development Trust, University of Aberdeen.

## Acknowledgements

The authors would like to thank all the participants who took time to annotate our reports and to attend the focus groups.

## Supplementary

### 1. MDT reports information

**Table 5.** Overview of simulated MDT reports and their use in the experiments.

|  | Cancer Type | Age | Gender | Clinical complexity | Used in the experiment |
| --- | --- | --- | --- | --- | --- |
| MDT 1 | Colorectal | 24 | M | Young patient with potential nodal involvement but candidate for potentially curative treatment | Sense checking/<br>Non-clinical annotation/ |
| MDT 2 | Colorectal | 94 | F | Elderly patient with bulky low rectal cancer. Only partial response to initial treatment. Pulmonary nodule on imaging. Potential surgical candidate | Sense checking/<br>Focus groups |
| MDT 3 | Colorectal | 65 | M | Comorbidities. - previous stroke. Metastatic disease detected and option of palliative chemotherapy | Sense checking/<br>Focus groups |
| MDT 4 | Colorectal | 63 | F | Case where the | Sense |
|  |  |  |  | clinical picture has evolved over time, with initially curative treatment, treatment for recurrence and now concern about potential recurrence in a lymph node | checking/<br>Non-clinical annotation/<br>Expert annotation/ |
| MDT 5 | Prostate | 71 | M | Not clinically complex – standard case. Different treatment options available to patient and also potential candidate for clinical trial | Sense checking/<br>Non-clinical annotation/<br>Clinical annotation/<br>Focus groups |
| MDT 6 | Prostate | 72 | M | High grade cancer with perineural invasion after prostatectomy. Several treatment options to be discussed with patient | Sense checking/<br>Non-clinical annotation/<br>Clinical annotation/<br>Focus groups |

### 2. Scenario prompt settings

Given the known context sensitivity of large language models, prompts were designed to reflect realistic clinical communication scenarios and were held constant within each evaluation task. Each prompt explicitly defined the role of the questioner, the clinical context, and the intended style of the response.

Clinician-facing scenarios: The model was instructed to assume the role of an experienced cancer consultant with 20 years of experience working within the UK National Health Service in Aberdeen. The prompt stated that the clinician had access to a multidisciplinary team (MDT) report and asked the model to either explain the report to a patient or give suggestions.

Patient-facing scenarios: The model was instructed to respond if the questioner was a cancer patient who had received their MDT report and found it difficult to understand and was seeking an explanation. The patient’s location (Aberdeen, UK) was specified to ensure responses were relevant to the healthcare context.

Each dialogue was limited to a single MDT report and a single role identity to avoid potential confounding effects of conversational history on subsequent model responses. No additional system prompts or conversational memory were provided.

### 3. Sense checking prompts examples

This section gives example prompts of the sense-checking experiments. We designed the following questions from multiple sources and input them to ChatGPT. Section 2.3.1 shows a detailed explanation.

- Hi, I’m 64, and I have been diagnosed with colorectal cancer. I was told I had an MDT meeting about me, and I’m pretty confused about the result. I asked my doctor to give me the MDT report. Feeling a bit down about the lack of information or support.
- Can my cancer be cured?
- I had resections before, but unfortunately, they recur. Why did that happen and is there something I should be careful about in daily life?
- What are the lymph nodes draining the right colon?
- How will I die?
- Should I take the chemotherapy?
- What is my prognosis?
- Why do my stroke and diabetes make treatment choices more challenging?
- How complex is my overall health?
- What does this mean “CT PET 22/2/22- 5 sites of PET Avidity. "
- How should I tell this to my family?
- Hi, I am a doctor and have a Prostate cancer patient report. Can you help me manage the information? You will be a very experienced prostate cancer specialist.
- Can you offer a detailed treatment plan with time for this patient, considering she lives in XXX?
- What should I do to release the patient’s anxiety and improve his life quality?

### 4. MDT and Questionnaire example

This is an example given to non-clinical people and clinicians during questionnaire annotation. The version presented to participants contained fabricated CHI numbers and patient and staff names. We anonymise sensitive information below to provide an understanding of the task and data.

**Dear colleague,**

Thank you for collaborating with us on our research project. We are interested in understanding how ChatGPT4 can handle complex patient documents. We have used mock reports from cancer multidisciplinary team (MDT) meetings, built to closely resemble real reports, but not using real patient data. We assumed the identity of the patient or doctor and asked ChatGPT a series of questions. We would like your help in identifying:

- Inaccurate responses
- Problems with the language used in the responses (e.g. too complex or technical, too ‘Americanised’, lack of emotional consideration)
- Other problems with the content of the response (e.g., seems reasonable but did not solve the problems)

We would like you to read the following examples and do three things:

1. **Annotate the ChatGPT output text (<u>not</u> the MDT report)** – you might want to print and annotate by hand or use highlighting/<u>underlining</u> or tracked changes and comments in Microsoft Word to highlight specific lines of text where you notice problems. Tell us what the problem is and please give any suggestions for how the ChatGPT response could be improved. (Please identify the problem type as classified above (inaccurate, language, other problems with content))
2. **Give the ChatGPT response an overall rating** – how well did you think ChatGPT handled the question in real-world cases?
3. **Provide some overall feedback** about the response in the comments box, telling us about things you liked or disliked about ChatGPT’s response, adding any details about any problems you found. What is missing?

Please send your completed responses to XXX. Thanks for your help!

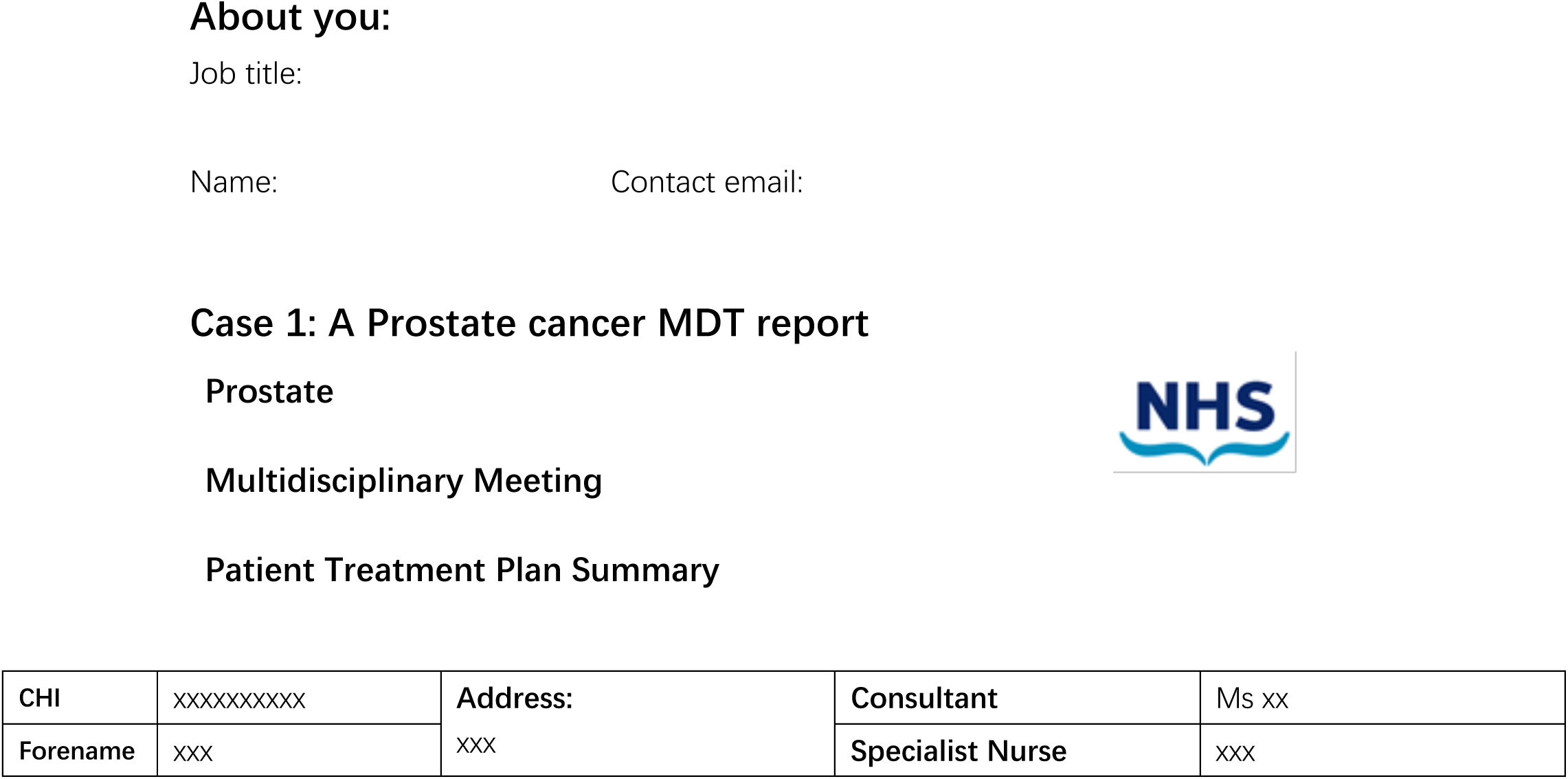
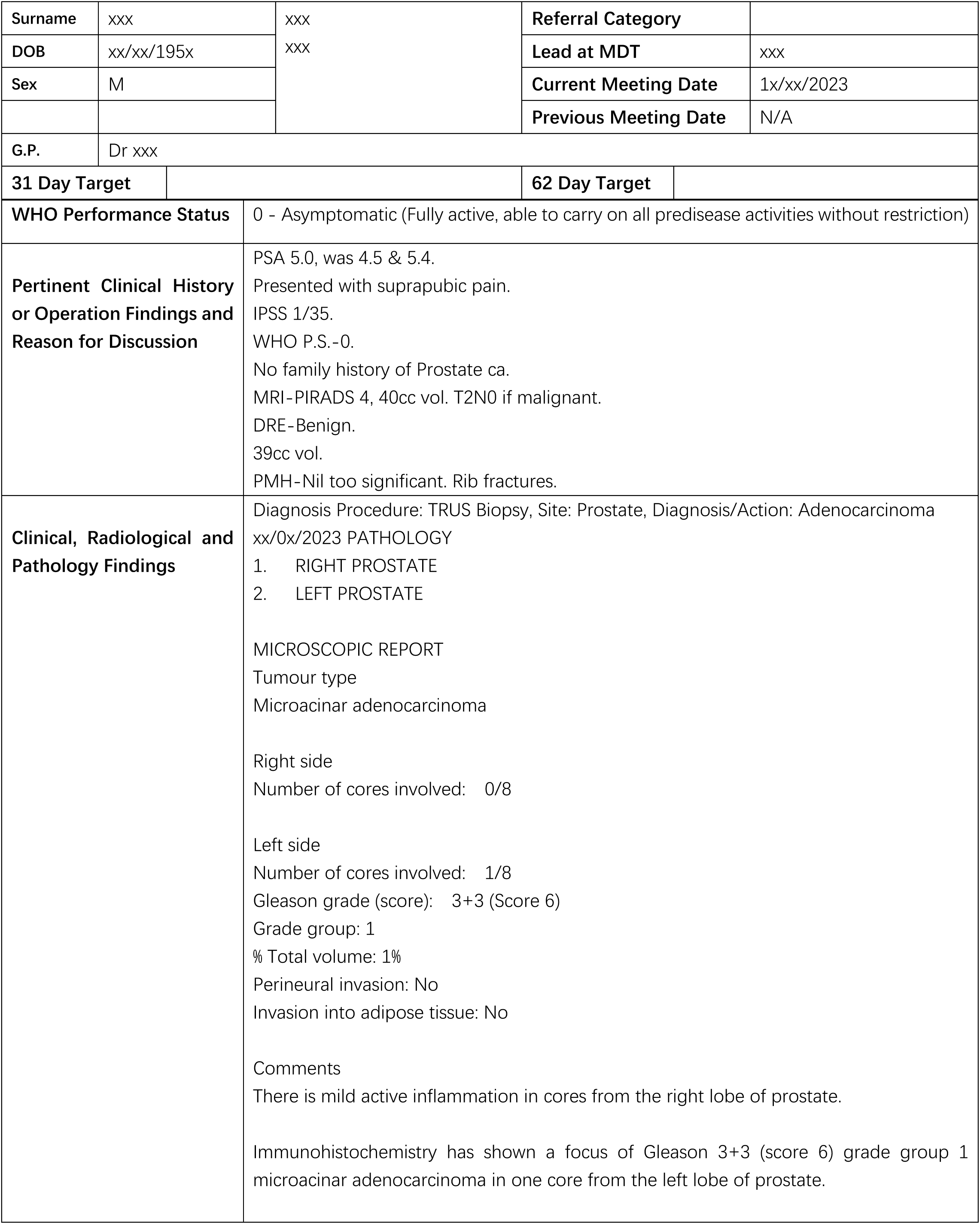
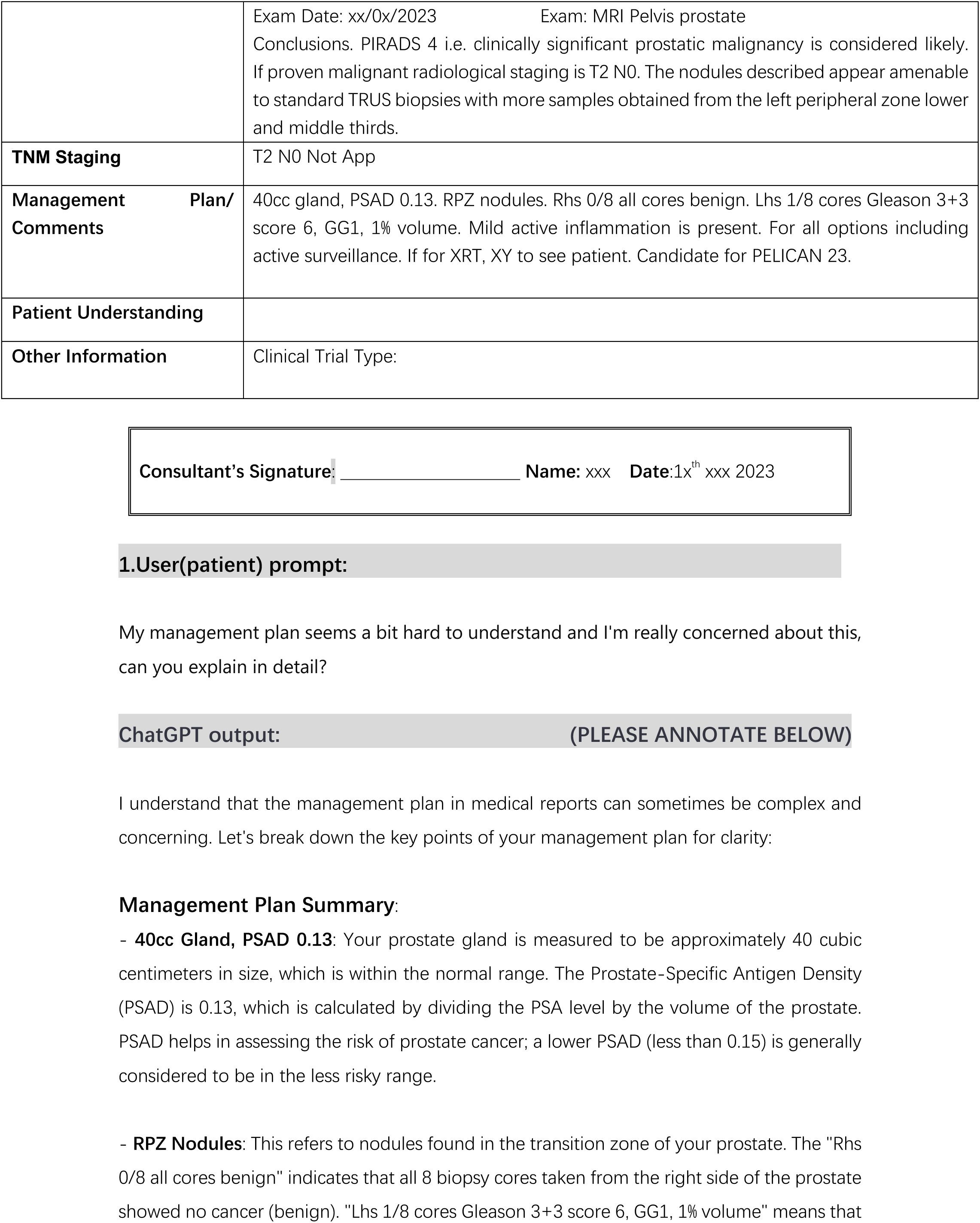
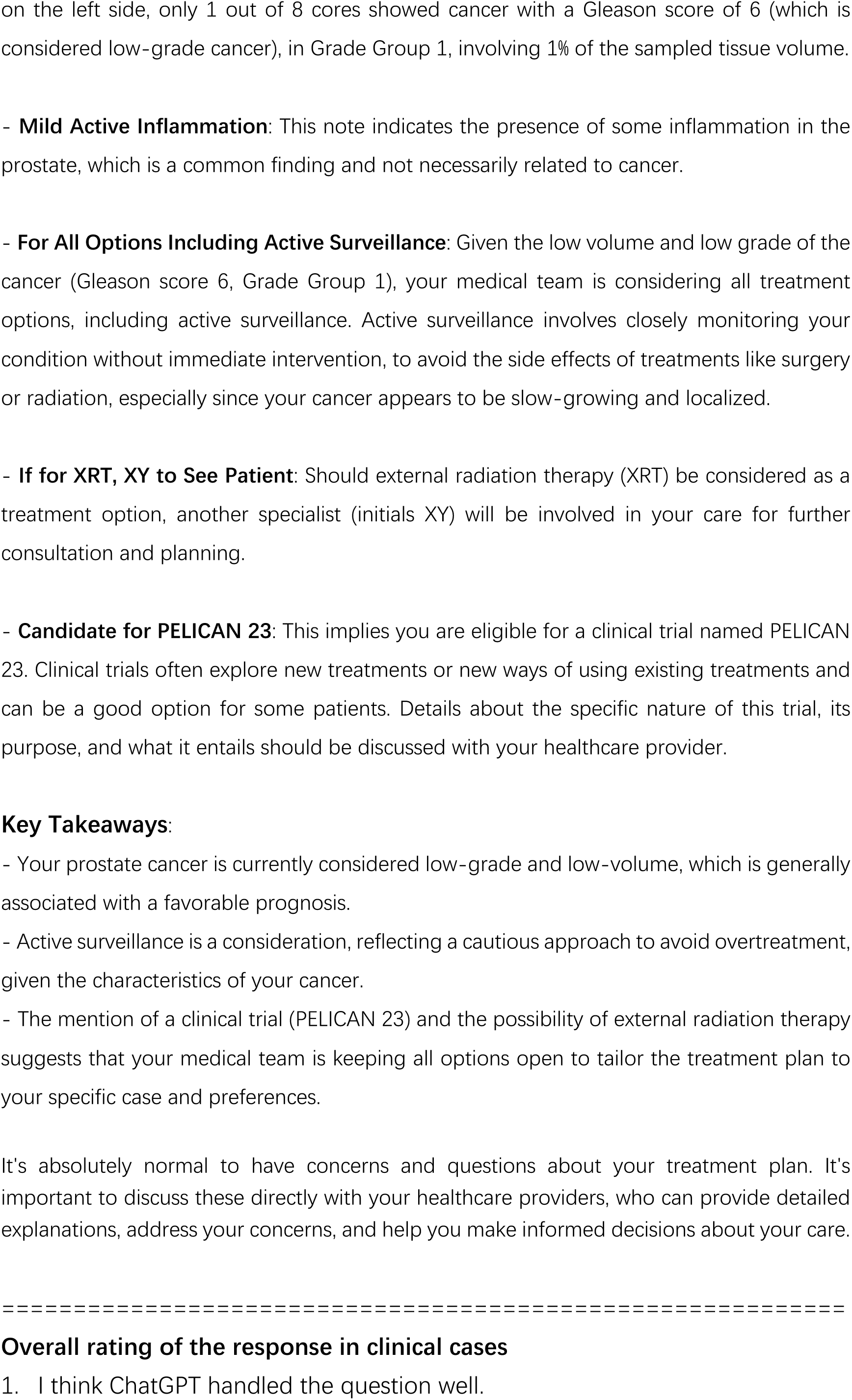
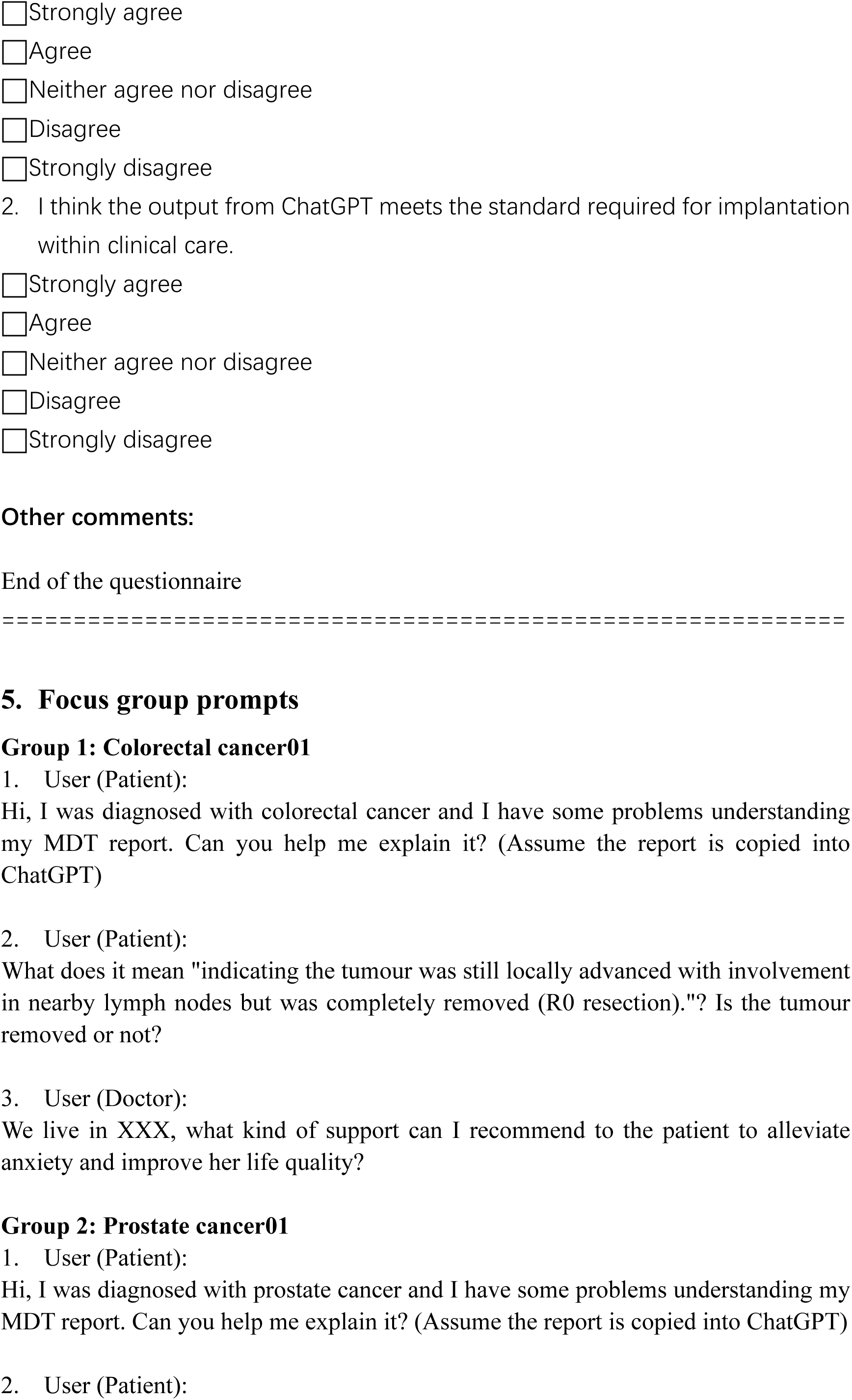
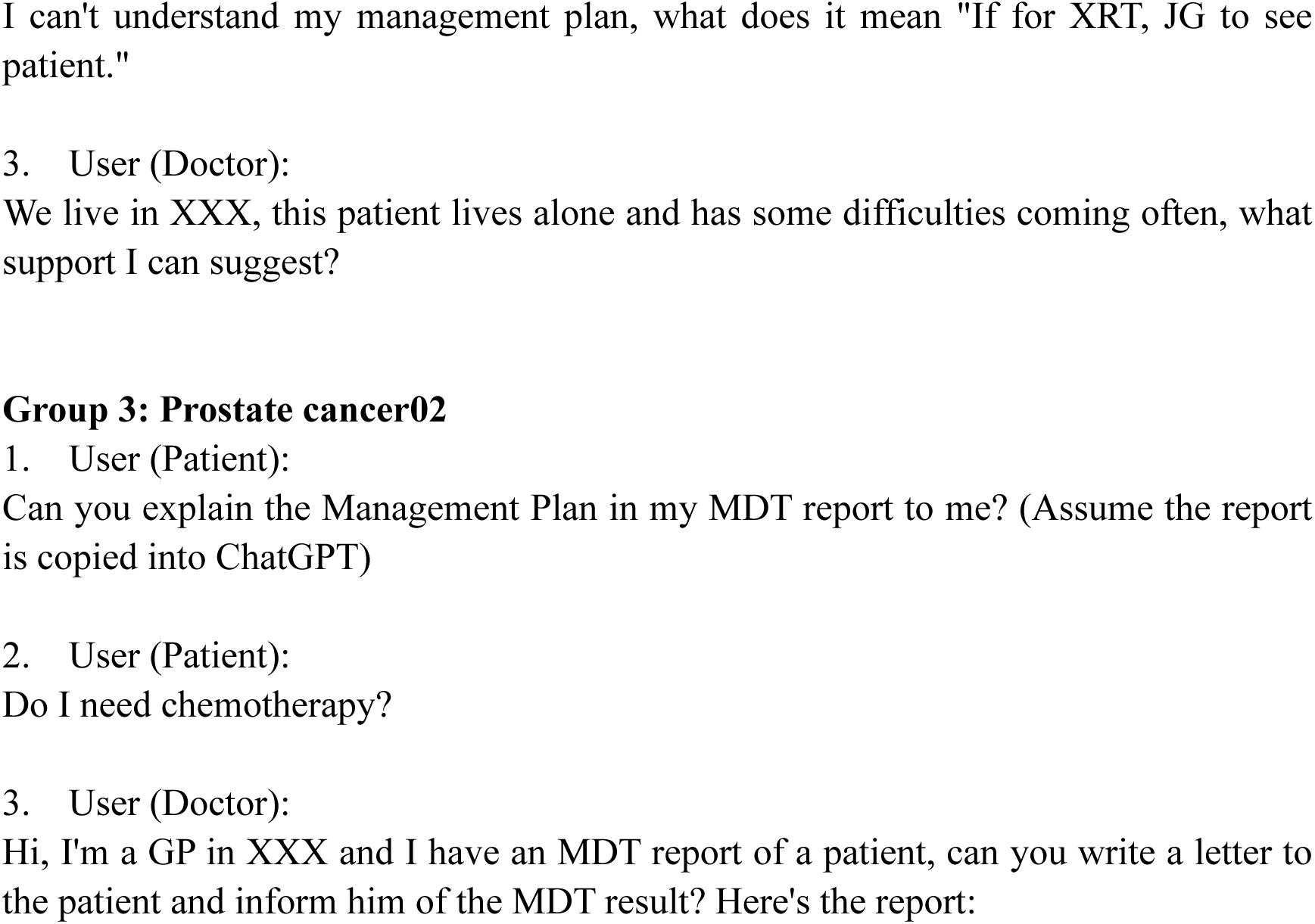

## Notes

### Competing Interest Statement

The authors have declared no competing interest.

### Author Declarations

The annotations experiment was approved by the School of Computing Science Ethics Committee, University of Aberdeen. Focus groups were conducted as part of a larger co-design event which was approved by Yorkshire and The Humber Leeds East Research Ethics Committee, reference 19/YH/0291 and received NHS R&D approval (NHS Grampian). All participants provided written informed consent.

## References

[1] D. C. Whiteman and L. F. Wilson, "The fractions of cancer attributable to modifiable factors: a global review," Cancer epidemiology, vol. 44, pp. 203–221, 2016.

[2] D. Lupton, "The digitally engaged patient: Self-monitoring and self-care in the digital health era," Social Theory & Health, vol. 11, no. 3, pp. 256–270, 2013.

[3] P. J. Fitzpatrick, "Improving health literacy using the power of digital communications to achieve better health outcomes for patients and practitioners," Frontiers in Digital Health, vol. 5, p. 1264780, 2023.

[4] B. Kane and S. Luz, "Information sharing at multidisciplinary medical team meetings," Group Decision and Negotiation, vol. 20, no. 4, pp. 437–464, 2011.

[5] M. Taberna et al., "The multidisciplinary team (MDT) approach and quality of care," Frontiers in oncology, vol. 10, p. 85, 2020.

[6] W. X. Zhao et al., "A survey of large language models," arXiv preprint arXiv:2303.18223, vol. 1, no. 2, 2023.

[7] OpenAI. "ChatGPT" https://chat.openai.com (accessed January 2023).

[8] R. Yang, T. F. Tan, W. Lu, A. J. Thirunavukarasu, D. S. W. Ting, and N. Liu, "Large language models in health care: Development, applications, and challenges," Health Care Science, vol. 2, no. 4, pp. 255–263, 2023.

[9] S. Shekar, P. Pataranutaporn, C. Sarabu, G. A. Cecchi, and P. Maes, "People Overtrust AI-Generated Medical Advice despite Low Accuracy," NEJM AI, vol. 2, no. 6, p. AIoa2300015, 2025.

[10] K. Singhal et al., "Towards expert-level medical question answering with large language models," Nature Medicine, 2023.

[11] S. Zhou et al., "The performance of large language model-powered chatbots compared to oncology physicians on colorectal cancer queries," International Journal of Surgery, vol. 110, no. 10, pp. 6509-6517, 2024.

[12] M. Sun, E. Reiter, L. Duncan, and R. Adam, "The role of natural language processing in improving cancer care: A scoping review with narrative synthesis," Artificial Intelligence in Medicine, p. 103227, 2025.

[13] Cancer.net. "Questions to Ask Your Health Care Team." https://www.cancer.net/navigating-cancer-care/diagnosing-cancer/questions-ask-your-health-care-team (accessed January 2023).

[14] National Cancer Institute. "Questions to Ask Your Doctor about Treatment." https://www.cancer.gov/about-cancer/treatment/questions (accessed January 2023).

[15] Macmillan Support. "Questions to ask your healthcare team." https://www.macmillan.org.uk/cancer-information-and-support/treatment/your-treatment-options/questions-to-ask-your-healthcare-team (accessed January 2023).

[16] Prostate cancer UK. "Prostate information and support." https://prostatecanceruk.org/prostate-information-and-support (accessed January 2023).

[17] Macmillan Support online community. "Bowel (colon and rectal) cancer forum." https://community.macmillan.org.uk/cancer_types/bowel-colon-rectum-cancer-forum?_gl=1*7oga1n*_up*MQ..&gclid=Cj0KCQiA35urBhDCARIsAOU7QwkVr0YKgpX9tr-9GtcAb-V5wdOAWQQmAI-rfweG9lH-T97dJMVDq7QaAlYnEALw_wcB&gclsrc=aw.ds (accessed January 2023).

[18] Cancer research uk forum. "Cancer Chat support forum." https://cancerchat.cancerresearchuk.org/f (accessed January 2023).

[19] C. A. McKim, "The value of mixed methods research: A mixed methods study," Journal of mixed methods research, vol. 11, no. 2, pp. 202–222, 2017.

[20] S. P. Wasti, P. Simkhada, E. R. van Teijlingen, B. Sathian, and I. Banerjee, "The growing importance of mixed-methods research in health," Nepal journal of epidemiology, vol. 12, no. 1, p. 1175, 2022.

[21] I. Etikan, S. A. Musa, and R. S. Alkassim, "Comparison of convenience sampling and purposive sampling," American journal of theoretical and applied statistics, vol. 5, no. 1, pp. 1–4, 2016.

[22] J. Golzar, S. Noor, and O. Tajik, "Convenience sampling," International Journal of Education & Language Studies, vol. 1, no. 2, pp. 72–77, 2022.

[23] H.-F. Hsieh and S. E. Shannon, "Three approaches to qualitative content analysis," Qualitative health research, vol. 15, no. 9, pp. 1277-1288, 2005.

[24] R. Adam et al., "Co-Design of the Structured Personalised Assessment for Reviews After Cancer (SPARC) Intervention," Health Expectations, vol. 28, no. 1, p. e70174, 2025.

[25] G. Terry, N. Hayfield, V. Clarke, and V. Braun, "Thematic analysis," The SAGE handbook of qualitative research in psychology, vol. 2, no. 17-37, p. 25, 2017.

[26] S. K. Ahmed et al., "Using thematic analysis in qualitative research," Journal of Medicine, Surgery, and Public Health, vol. 6, p. 100198, 2025.

[27] S. Frenda et al., "Perspectivist approaches to natural language processing: a survey," Language Resources and Evaluation, pp. 1–28, 2024.

[28] A. N. Uma, T. Fornaciari, D. Hovy, S. Paun, B. Plank, and M. Poesio, "Learning from disagreement: A survey," Journal of Artificial Intelligence Research, vol. 72, pp. 1385-1470, 2021.

[29] E. Reiter, Natural Language Generation. Springer, 2025.

[30] F. Moramarco, "Evaluation of medical note generation systems" PhD Thesis, Aberdeen University, 2024.

[31] M. J. Duggan et al., "Clinician experiences with ambient scribe technology to assist with documentation burden and efficiency," JAMA Network Open, vol. 8, no. 2, pp. e2460637-e2460637, 2025.

[32] J. M. Choo et al., "Conversational artificial intelligence (chatGPT™) in the management of complex colorectal cancer patients: early experience," ANZ journal of surgery, vol. 94, no. 3, pp. 356–361, 2024.

[33] A. Derton et al., "Natural language processing methods to empirically explore social contexts and needs in cancer patient notes," JCO Clinical Cancer Informatics, vol. 7, p. e2200196, 2023.

[34] S. Lukac et al., "Evaluating ChatGPT as an adjunct for the multidisciplinary tumor board decision-making in primary breast cancer cases," Archives of Gynecology and Obstetrics, vol. 308, no. 6, pp. 1831-1844, 2023.

[35] J. Vela Ulloa, S. King Valenzuela, C. Riquoir Altamirano, and G. Urrejola Schmied, "Artificial intelligence-based decision-making: can ChatGPT replace a multidisciplinary tumour board?," British Journal of Surgery, vol. 110, no. 11, pp. 1543-1544, 2023.

[36] J. Haemmerli, et al., "ChatGPT in glioma adjuvant therapy decision making: ready to assume the role of a doctor in the tumour board?," BMJ health & care informatics, vol. 30, no. 1, p. e100775, 2023.

[37] V. Sorin et al., "Large language model (ChatGPT) as a support tool for breast tumor board," NPJ Breast Cancer, vol. 9, no. 1, p. 44, 2023.

[38] M. L. Specchia et al., "The impact of tumor board on cancer care: evidence from an umbrella review," BMC health services research, vol. 20, no. 1, p. 73, 2020.

[39] M. A. Fink et al., "Potential of ChatGPT and GPT-4 for data mining of free-text CT reports on lung cancer," Radiology, vol. 308, no. 3, p. e231362, 2023.

[40] D. Hu, B. Liu, X. Zhu, X. Lu, and N. Wu, "Zero-shot information extraction from radiological reports using ChatGPT," International Journal of Medical Informatics, vol. 183, p. 105321, 2024.

[41] E. M. Chung, S. C. Zhang, A. T. Nguyen, K. M. Atkins, H. M. Sandler, and M. Kamrava, "Feasibility and acceptability of ChatGPT generated radiology report summaries for cancer patients," Digital health, vol. 9, p. 20552076231221620, 2023.

[42] K. Jeblick, et al., "ChatGPT makes medicine easy to swallow: an exploratory case study on simplified radiology reports," European radiology, vol. 34, no. 5, pp. 2817-2825, 2024.

[43] Y. Nakamura et al., "ChatGPT for automating lung cancer staging: feasibility study on open radiology report dataset," medRxiv, p. 2023.12. 11.23299107, 2023.

[44] T. H. Kung, et al., "Performance of ChatGPT on USMLE: potential for AI-assisted medical education using large language models," PLoS digital health, vol. 2, no. 2, p. e0000198, 2023.

[45] A. Gilson et al., "How does ChatGPT perform on the United States medical licensing examination? The implications of large language models for medical education and knowledge assessment," JMIR Medical Education, vol. 9, no. 1, p. e45312, 2023.

[46] J. Kasai, Y. Kasai, K. Sakaguchi, Y. Yamada, and D. Radev, "Evaluating gpt-4 and chatgpt on japanese medical licensing examinations," arXiv preprint arXiv:2303.18027, 2023.

[47] A. Madrid-García et al., "Harnessing ChatGPT and GPT-4 for evaluating the rheumatology questions of the Spanish access exam to specialized medical training," Scientific Reports, vol. 13, no. 1, p. 22129, 2023.

[48] B. Koopman and G. Zuccon, "Dr ChatGPT tell me what I want to hear: How different prompts impact health answer correctness," in Proceedings of the 2023 conference on empirical methods in natural language processing, 2023, pp. 15012-15022.

[49] L. Mehnen, S. Gruarin, M. Vasileva, and B. Knapp, "ChatGPT as a medical doctor? A diagnostic accuracy study on common and rare diseases," MedRxiv, p. 2023.04. 20.23288859, 2023.

[50] L. Tang et al., "Evaluating large language models on medical evidence summarization," NPJ digital medicine, vol. 6, no. 1, p. 158, 2023.

[51] S. Balloccu, P. Schmidtová, M. Lango, and O. Dušek, "Leak, cheat, repeat: Data contamination and evaluation malpractices in closed-source LLMs," arXiv preprint arXiv:2402.03927, 2024.

[52] T. Tu et al., "Towards conversational diagnostic AI," arXiv preprint arXiv:2401.05654, 2024.

[53] S. Chen et al., "Use of artificial intelligence chatbots for cancer treatment information," JAMA oncology, vol. 9, no. 10, pp. 1459-1462, 2023.

[54] S. Chen et al., "The effect of using a large language model to respond to patient messages," The Lancet Digital Health, vol. 6, no. 6, pp. e379-e381, 2024.

[55] A. Tariq et al., "Domain-specific llm development and evaluation–a case-study for prostate cancer," medRxiv, p. 2024.03. 15.24304362, 2024.

